# A SYSTEMATIC RE-ANALYSIS OF COPY NUMBER LOSSES OF UNCERTAIN CLINICAL SIGNIFICANCE

**DOI:** 10.1101/2023.07.02.23292123

**Authors:** George J Burghel, Jamie M Ellingford, Ronnie Wright, Lauren Bradford, Jake Miller, Christopher Watt, Jonathan Edgerley, Farah Naeem, Siddharth Banka

## Abstract

**Background:** Re-analysis of whole exome/genome data improves diagnostic yield. However, the value of re-analysis of clinical array comparative genomic hybridisation (aCGH) data has never been investigated. Case-by-case re-analysis is impractical in busy diagnostic laboratories.

**Methods and Results:** We harmonised historical post-natal clinical aCGH results from ∼16,000 patients tested via our diagnostic laboratory over ∼7 years with current clinical guidance. This led to identification of 33,857 benign, 2,173 class 3, and 979 pathogenic copy number losses (CNLs). We found benign CNLs to be significantly less likely to encompass haploinsufficient genes compared to the pathogenic or class 3 CNLs in our database. Using this observation, we developed a re-analysis pipeline (using up-to-date disease association data and haploinsufficiency scores) and shortlisted 207 class 3 CNLs encompassing at least one autosomal dominant disease-gene associated with haploinsufficiency or loss-of-function mechanism. Clinical scientist review led to reclassification of 7.2% shortlisted class 3 CNLs as pathogenic or likely pathogenic. This included first cases of CNV-mediated disease for some genes where all previously described cases involved only point variants. Interestingly, some CNLs could not be re-classified because the phenotypes of patients with CNLs seemed distinct from the known clinical features resulting from point variants, thus raising questions about accepted underlying disease mechanisms. Several potential novel disease-genes were identified that would need further validation.

**Conclusions:** Re-analysis of clinical aCGH data increases diagnostic yield and demonstrates their research value. In future, the aCGH reanalysis program should be expanded to include other copy number variant types.

## Introduction

Copy number variants (CNVs) are a major cause of developmental disorders (Coe et al., 2014; Cooper et al., 2011; Mefford et al., 2009). Pathogenic CNVs result in diverse phenotypes and can have significant medical and socioeconomic impact (Burghel et al., 2019). Array comparative genomic hybridisation (aCGH), has now been used for more than a decade as a first-line diagnostic test for children with neurodevelopmental disorders (Miller et al., 2010). The diagnostic yield of aCGH in diagnostic laboratories is ∼12% (Miller et al., 2010). Clinical interpretation of aCGH data frequently results in classification of variants as having uncertain significance (VUS) (Riggs et al., 2020). Accurate resolution of variants of uncertain significance (VUS) is a major issue in the field of clinical genomics (Hoffman-Andrews, 2017). Deeper interrogation of CNV data can improve clinical interpretation, lead to discovery of new genetic disorders and provide novel insights into fundamental biology and disease mechanisms (Banka et al., 2015; Kasher et al., 2016; Tinker et al., 2020). Previous studies have shown that periodic reassessment of whole exome sequencing (WES) or whole genome sequencing (WGS) data helps in the reclassification of VUS as (likely) benign or pathogenic, and improves diagnostic yield (Tan et al., 2020), (Hiatt et al., 2018; Salfati et al., 2019; Sun et al., 2019) (Bruel et al., 2019; Costain et al., 2018; Das et al., 2014) (Wright et al., 2018). However, so far there have not been many studies of the value of reinterpretation of aCGH data. Re-analysis of thousands of class 3 CNVs detected in diagnostic laboratories over a period of years can be challenging and time-consuming. Case-by-case analysis requires significant resources and is challenging. We, therefore, decided to devise an approach for efficient systematic reanalysis of class 3 copy number losses detected in our diagnostic laboratory.

## Methods

### CNV classification and annotation

As previously described, a departmental database was curated from pseudo-anonymised postnatal clinical array-CGH testing performed at MCGM between 2010 and 2017 (Burghel et al., 2020). Copy number losses (CNLs) were divided into three groups based on reported clinical classification; benign (classes 1 and 2), VUSs (class 3) and pathogenic (classes 4 and 5). Bedtools intersect was used to identify regions that were unique to pathogenic variants and those unique to VUS events. CNLs of >4.8Mb were excluded from subsequent analysis.

### Gene-centric analysis and shortlisting

Genes overlapping with any of the three CNL classification groups (minimal CNL coordinates (Human genome build hg19)) were identified based on genomic location. Genes were annotated with Online Mendelian Inheritance in Man (OMIM) number, associated OMIM disorder and mode of inheritance (if available/applicable); the Development Disorder Genotype - Phenotype Database (DDG2P); and dosage sensitivity scores where available (probability of being loss-of-function (LoF) intolerant (pLI), Residual Variation Intolerance Score (RVIS) and the Decipher haploinsufficiency (D-HI)) (Huang et al., 2010; Lek et al., 2016; Lessard et al., 2016; Petrovski et al., 2013).

All genes within cases that have pathogenic CNLs alongside the class 3 CNLs were excluded. Genes within common CNLs (occurring in ≥100 cases) were also excluded, because these common CNLs are likely to be either artefactual or (likely) benign (Figure 1). Genes were shortlisted for further analysis if they were associated with an OMIM syndrome with autosomal dominant inheritance and/or a loss of function consequence as defined by DDG2P. Genes were further shortlisted if they were predicted to be haploinsufficient with pLI score ≥0.9 and RVIS score ≤0.01 (Figure 1).

**Figure 1:**
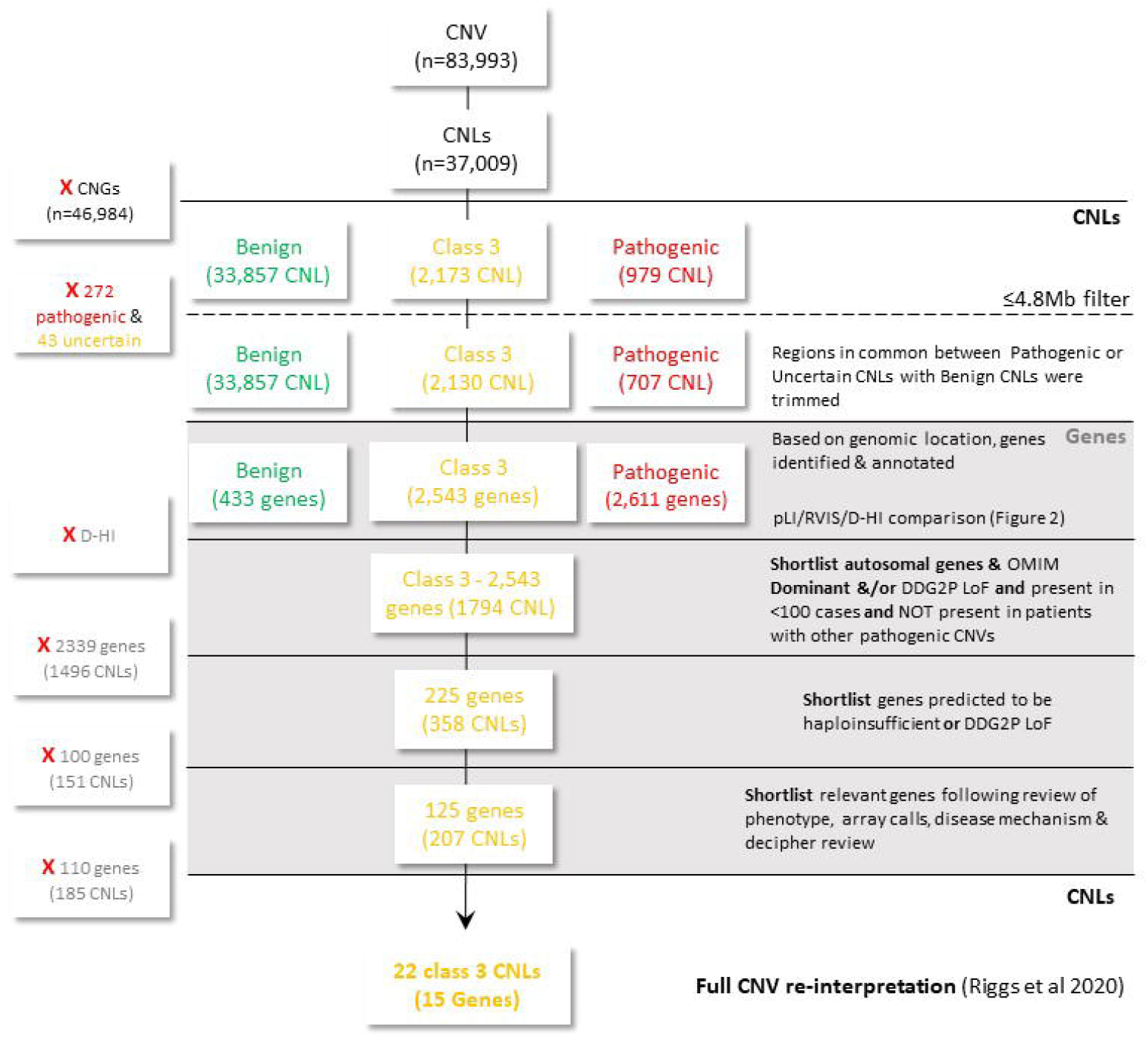
Systematic CNL analysis.

### Statistical analysis

Chi squared tests of independence and trend were performed and statistical significance was set at p-value <0.05. The tests were performed to examine the associations between three haploinsufficiency prediction scores and the clinical classification.

### Analysis of shortlisted genes and re-classification

Genes were further shortlisted manually by a Clinical Scientist. This was based on relevance of associated phenotype (relevant phenotypes included developmental delays, congenital anomalies, autistic spectrum disorders etc), mechanism of disease (loss of function), manual review of the array calls (where possible) and any similar CNVs on Decipher. Any CNLs that included shortlisted genes after this manual review had a full clinical interpretation following local procedures, the recent ClinGen/ACMG guidance and the ClinGen CNV pathogenicity calculator (https://cnvcalc.clinicalgenome.org/cnvcalc/) (Riggs et al., 2020).

## Results

### Haploinsufficiency scores correlate with clinical pathogenicity classification of CNLs

We identified 37,009 CNLs from >16,000 postnatal clinical array-CGHs performed at the MCGM between 2010 and 2017. Of these 33,857 CNLs were classed as benign or likely benign; 2,173 were classed as VUS; and 979 were classed as likely pathogenic or pathogenic (Figure 1). From subsequent analysis, 3,152 CNLs of 4.8Mb or more were excluded. The CNLs were distributed across the genome (Supplementary Figure S1). Trimming of regions shared with benign CNLs from pathogenic and class 3 VUS identified 2,611 genes within regions affected by only pathogenic CNLs and 2,543 genes within regions affected by class 3 CNLs and 433 genes in benign CNLs.

Depending on the metric used for haploinsufficiency, we found 78-96% of pathogenic/likely pathogenic CNLs to include at least one haploinsufficiency intolerant gene, while 7-49% of benign/likely benign CNLs were found to have at least one haplosinufficiency intolerant gene. This difference was statistically significant across the three haploinsufficiency scores; pLI ∼84% (pathogenic) compared to ∼7% (benign); RVIS ∼96% (pathogenic) versus ∼13% (benign); and D-HI ∼78% (pathogenic) compared to ∼49% (benign) (P=<0.00001 X2-Test) (Figure 2). In comparison, 34-55% of 3 CNLs were found to have at least one haploinsufficiency intolerant gene (Table 1). These differences were significant for class 3 CNLs but only for pLI (∼34% (class 3) vs ∼7% (benign)) and RVIS (∼54% (class 3) vs ∼13% (benign) (P=<0.00001 X2-Test). In contrast, according to the D-HI score, benign CNLs were more likely to have ≥1 haploinsufficiency intolerant gene (∼49%) compared to class 3 CNLs (∼35%) (P=<0.00001 X2-Test) (Figure 2).

**Table 1.** CNLs with at least one haploinsufficient gene

| CNL class (Total No of CNLs) | Unique CNLs (n) | Predicted Haploinsufficient genes |  |  |
| --- | --- | --- | --- | --- |
|  |  | pLI (%) | RVIS (%) | D-HI (%) |
| Pathogenic (707) | 410 | 345 (84.1) | 394 (96.1) | 321 (78.3) |
| Class 3 (2,130) | 1,489 | 509 (34.2) | 812 (54.5) | 516 (34.7) |
| Benign (3,3857) | 1,314 | 95 (7.2) | 174 (13.2) | 643 (48.9) |

**Figure 2:**
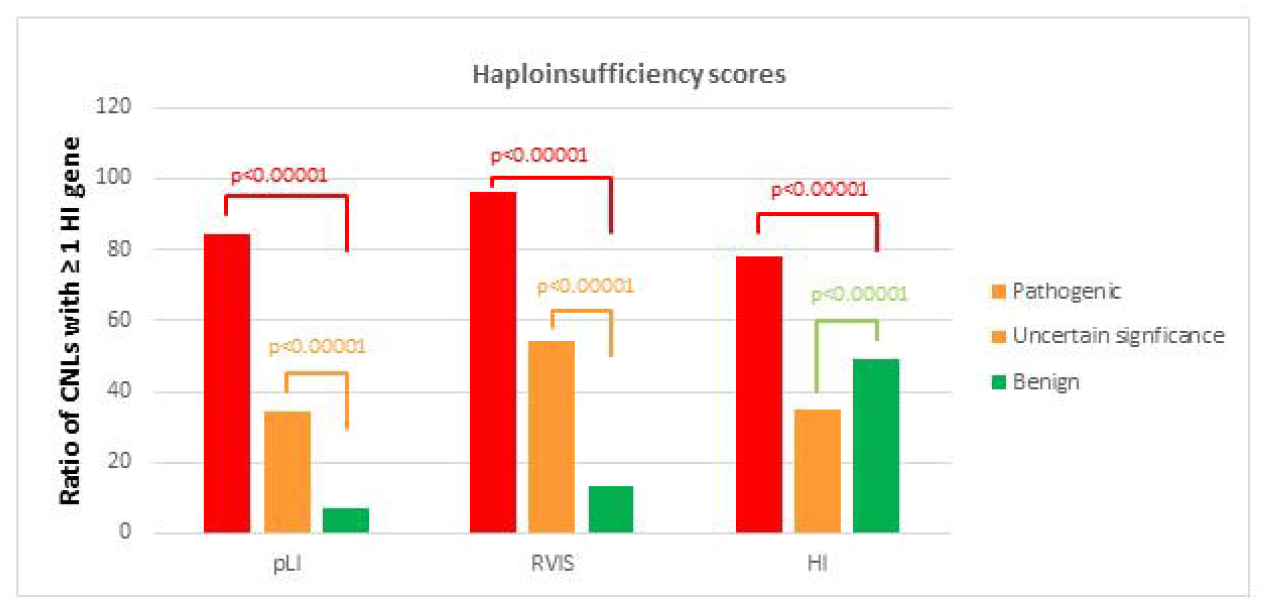
Haploinsufficiency scores enrichment. Comparison of CNLs with ≥ 1 Haploinsufficient genes across the 3 CNL categories (Pathogenic, uncertain significance and benign). Please note that ratios are presented here, while X2-Test p-value is based on total numbers. Details Supplementary Table S1.

Overall, out of all the genes deleted only in pathogenic CNLs, ∼19% were considered to be haploinsufficeint by pLI and ∼21% by RVIS. Similar patterns were observed for class 3 CNLs (pLI ∼19% and RVIS ∼24%). Using pLI and RVIS, both pathogenic and class 3 CNLs had higher levels of haploinsufficient genes compared to benign CNLs (∼12% (PLI) and ∼16% (RVIS)) (Supplementary Table S1). In contrast D-HI seems to predict more haploinsufficient genes within benign CNLs (∼79%) compared to pathogenic CNLs (∼55%) and class 3 CNLs (∼51%) (Supplementary Table S1). Due to the over-prediction of haploinsufficient genes using D-HI within benign CNLs, this score was excluded from further analysis.

Collectively, the enrichment of haploinsufficient genes as predicted by pLI and RVIS in pathogenic CNLs indicated that these are reliable scores to identify haploinsfficient genes within class 3 CNLs.

### Reclassification of copy number losses of uncertain significance to pathogenic

A total of 2,543 genes (1,794 class 3 CNLs) were annotated for information from OMIM, DDG2P in addition to haploinsufficiency scores (pLI and RVIS) (Figure 1). We included autosomal genes with dominant OMIM disease association and/or LoF mutation consequence by DDG2P, present in <100 cases and absent in patients who had other pathogenic CNVs. This resulted in a shortlist of 225 genes (in 358 CNLs). Following that, genes were further shortlisted based on being haploinsufficient as predicted by both pLI (score ≥0.9) and RVIS (score ≤0.01) scores but also including DDG2P mutation consequence of LoF. This resulted in a list of 125 genes (207 CNLs) (Figure 1, Supplementary Table S2).

This list was then assessed by clinical scientists and this included; review of original aCGH data (where possible) to exclude any likely-artefactual CNLs and non-coding CNLs; examining phenotypic information of associated syndrome(s) to include relevant phenotype (e.g. developmental delays, congenital anomalies, autistic spectrum disorders etc) and exclude any non-relevant/late age of onset phenotypes (such as cancer) and likely mechanism of pathogenicity (LoF vs gain of function (GoF)) in addition to similar and overlapping Decipher cases. The clinical scientist review resulted in a priority shortlist of 15 genes (22 CNLs).

The 22 CNLs had a full CNV interpretation as per the local procedures and latest ACMG/ClinGen criteria (Riggs et al., 2020). This resulted in a total of 15 CNL reclassifications, all of which were due to genes and disease association (or disease-causing mechanism) described post the identification and the original classification and reporting of the CNV (Table 2).

**Table 2.**
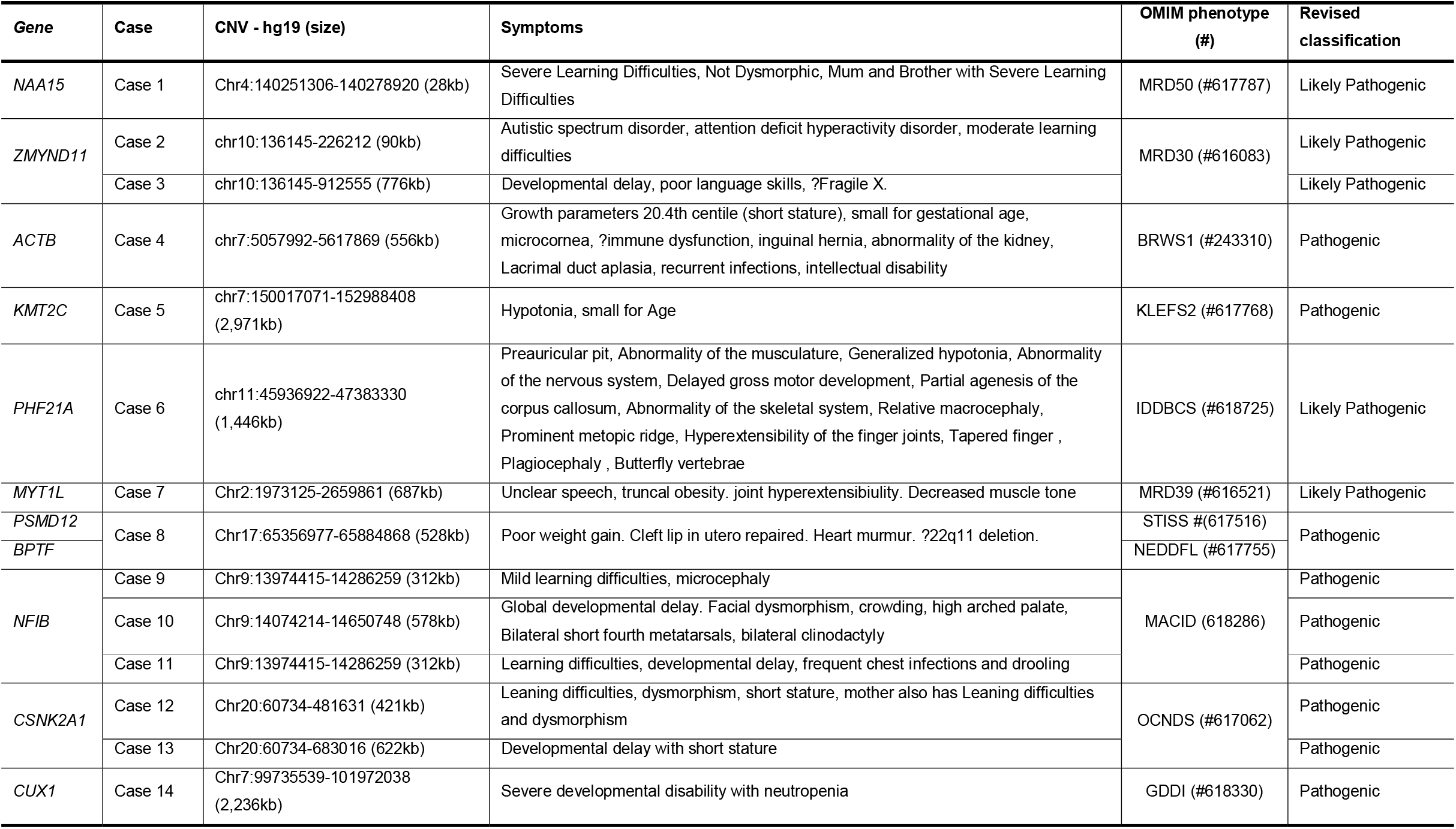
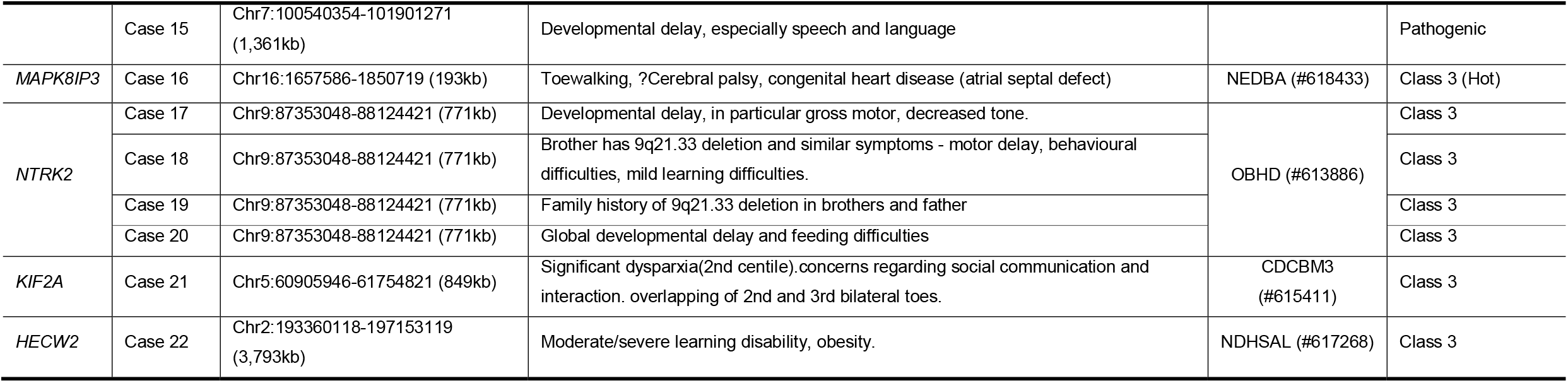
Prioritised Genes and the associated CNLs that have undergone reclassification

| **Gene** | **Case** | **CNV - hg19 (size)** | **Symptoms** | **OMIM phenotype (#)** | **Revised classification** |
| --- | --- | --- | --- | --- | --- |
| *NAA15* | Case 1 | Chr4:140251306-140278920 (28kb) | Severe Learning Difficulties, Not Dysmorphic, Mum and Brother with Severe Learning Difficulties | MRD50 (#617787) | Likely Pathogenic |
| *ZMYND11* | Case 2 | chr10:136145-226212 (90kb) | Autistic spectrum disorder, attention deficit hyperactivity disorder, moderate learning difficulties | MRD30 (#616083) | Likely Pathogenic |
|  | Case 3 | chr10:136145-912555 (776kb) | Developmental delay, poor language skills, ?Fragile X. |  | Likely Pathogenic |
| *ACTB* | Case 4 | chr7:5057992-5617869 (556kb) | Growth parameters 20.4th centile (short stature), small for gestational age, microcornea, ?immune dysfunction, inguinal hernia, abnormality of the kidney, Lacrimal duct aplasia, recurrent infections, intellectual disability | BRWS1 (#243310) | Pathogenic |
| *KMT2C* | Case 5 | chr7:150017071-152988408 (2,971kb) | Hypotonia, small for Age | KLEFS2 (#617768) | Pathogenic |
| *PHF21A* | Case 6 | chr11:45936922-47383330 (1,446kb) | Preauricular pit, Abnormality of the musculature, Generalized hypotonia, Abnormality of the nervous system, Delayed gross motor development, Partial agenesis of the corpus callosum, Abnormality of the skeletal system, Relative macrocephaly, Prominent metopic ridge, Hyperextensibility of the finger joints, Tapered finger , Plagiocephaly , Butterfly vertebrae | IDDBCS (#618725) | Likely Pathogenic |
| *MYT1L* | Case 7 | Chr2:1973125-2659861 (687kb) | Unclear speech, truncal obesity. joint hyperextensibility. Decreased muscle tone | MRD39 (#616521) | Likely Pathogenic |
| *PSMD12* | Case 8 | Chr17:65356977-65884868 (528kb) | Poor weight gain. Cleft lip in utero repaired. Heart murmur. ?22q11 deletion. | STISS # (617516) | Pathogenic |
| *BPTF* |  |  |  | NEDDFL (#617755) |  |
| *NFIB* | Case 9 | Chr9:13974415-14286259 (312kb) | Mild learning difficulties, microcephaly | MACID (618286) | Pathogenic |
|  | Case 10 | Chr9:14074214-14650748 (578kb) | Global developmental delay. Facial dysmorphism, crowding, high arched palate, Bilateral short fourth metatarsals, bilateral clinodactyly |  | Pathogenic |
|  | Case 11 | Chr9:13974415-14286259 (312kb) | Learning difficulties, developmental delay, frequent chest infections and drooling |  | Pathogenic |
| *CSNK2A1* | Case 12 | Chr20:60734-481631 (421kb) | Leaning difficulties, dysmorphism, short stature, mother also has Leaning difficulties and dysmorphism | OCNDS (#617062) | Pathogenic |
|  | Case 13 | Chr20:60734-683016 (622kb) | Developmental delay with short stature |  | Pathogenic |
| *CUX1* | Case 14 | Chr7:99735539-101972038 (2,236kb) | Severe developmental disability with neutropenia | GDDI (#618330) | Pathogenic |
|  | Case 15 | Chr7:100540354-101901271<br>(1,361kb) | Developmental delay, especially speech and language |  | Pathogenic |
| *MAPK8IP3* | Case 16 | Chr16:1657586-1850719 (193kb) | Toewalking, ?Cerebral palsy, congenital heart disease (atrial septal defect) | NEDBA (#618433) | Class 3 (Hot) |
| *NTRK2* | Case 17 | Chr9:87353048-88124421 (771kb) | Developmental delay, in particular gross motor, decreased tone. | OBHD (#613886) | Class 3 |
|  | Case 18 | Chr9:87353048-88124421 (771kb) | Brother has 9q21.33 deletion and similar symptoms - motor delay, behavioural difficulties, mild learning difficulties. |  | Class 3 |
|  | Case 19 | Chr9:87353048-88124421 (771kb) | Family history of 9q21.33 deletion in brothers and father |  | Class 3 |
|  | Case 20 | Chr9:87353048-88124421 (771kb) | Global developmental delay and feeding difficulties |  | Class 3 |
| *KIF2A* | Case 21 | Chr5:60905946-61754821 (849kb) | Significant dyspraxia(2nd centile).concerns regarding social communication and interaction. overlapping of 2nd and 3rd bilateral toes. | CDCBM3<br>(#615411) | Class 3 |
| *HECW2* | Case 22 | Chr2:193360118-197153119<br>(3,793kb) | Moderate/severe learning disability, obesity. | NDHSAL (#617268) | Class 3 |

Collectively, this strategy enabled reclassification rate is 7.2% of the shortlisted CNLs (207 CNLs). Notably, this was∼0.7% of the starting cohort of 2,173 CNLs.

## Discussion

Although CNV analysis has been a first-tier genomic diagnostic test for more than decade, utility of re-analysis of its data has not been studied. Most diagnostic laboratories revisit class 3 variants reactively, for example if this was indicated by a clinician or another similar variant was identified in another patient (El Mecky et al., 2019). Here, we devised an approach to efficiently facilitate systematic re-analysis of class 3 CNLs at a large-scale.

We used updated OMIM annotations in addition to haploinsufficiency scores to short list class 3 CNLs of interest. Our haploinsufficiency score landscape was akin to previously published data. Our analysis of pathogenic CNLs (after excluding overlap with benign CNLs) have shown that 19.4% of genes within these regions are predicted to be haploinsufficient by pLI scores while only 12.4% in the benign CNLs. This is comparable to the results from an analysis performed on CNLs obtained from the database of structural variants (dbVar) (3,269 pathogenic CNLs and 3,699 benign CNLs), whereby they found that 19.5% of the genes exclusively observed in pathogenic CNV regions to be haploinsufficient (by pLI) compared to 12% in benign regions. RVIS scores have shown similar patterns (21.1% for pathogenic CNLs and 15.9% for benign ones). On the other hand, D-HI scores indicate haploinsufficient genes to be more prevalent in benign CNLs (79%) compared to pathogenic CNLs (55%). D-HI scores were predicted based on an evolutionary and functional similarities to ∼300 established haploinsufficient genes (Huang et al., 2010). In contrast, pLI and RVIS scores were calculated for all genes based on comparing observed protein truncating variants with expected ones based on gene size (Lek et al., 2016; Petrovski et al., 2013). These differences in computing these scores could account for the better performance of pLI and RVIS in comparison to D-HI. Based on this, we excluded the D-HI scores from further analyses and confirmed that pLI and RVIS were robust scores for predicting haploinsufficient genes in our cohort. Further comparison of haplosinsufficiency scores has also shown enrichment of haploinsufficient genes in class 3 CNLs compared to benign ones. This indicated the increased likelihood of the presence of pathogenic deletions amongst the variants of uncertain significance (class 3 CNLs).

We then followed a systematic filtering and shortlisting process (Figure 1) that resulted in a short list of 125 genes. These genes were interrogated by trained clinical scientists reviewing associated phenotypes, disease mechanism and array calls. This resulted in a final short list of 15 genes (22 CNLs) that warranted full reinterpretation according to the latest guidance (Riggs et al., 2020). This resulted in 15 CNLs (11 genes) reclassifications to (likely) pathogenic. All the reclassifications were due to genes and disease association described post the identification and reporting of the class 3 CNLs, apart from ACTB, whereby disease association was known prior to the class 3 CNL reporting, however, the loss of function disease mechanism was not confirmed until 2017 (post class 3 CNL reporting). The final reclassification rate was ∼0.7% of the total number of class 3 CNLs identified at the start of this study but importantly ∼7.2% of the shortlisted 207 CNLs.

The reclassification rate is lower than reported rates of reclassification from most NGS based studies (multi-gene panels, WES and WGS) (Bruel et al., 2019; Costain et al., 2018; Das et al., 2014; Hiatt et al., 2018; Sun et al., 2019; Wright et al., 2018). However, there are no similar studies from aCGH data for comparison. This lower rate could be attributed to the more targeted nature of aCGH compared to exome and genome analysis. Apart from the inherent limitation of the aCGH design, this study has several limitations that may impact reclassification rates. The shortlisting of genes was mainly reliant on OMIM phenotype associations and haploinsufficient score predictions. The OMIM phenotype associations could miss some interesting genes with recent publications compared to literature searches, however, the latter is not suitable for automating the process. Haploinsufficient score predictions were used as proxy for haploinsufficiency and although are reasonably predictive they are not accurate for all genes as we have recently demonstrated (Tinker et al., 2020). Moreover, we haven’t attempted the identification of contiguous gene disorders nor reclassification of class 3 CNLs that may disrupt regulatory regions and TADs. We also did not attempt the reclassification of class 3 CNVs into benign categories using updated population databases. Finally, we have only focussed on CNLs and not copy number gains (CNGs). All of these extra analyses could potentially increase the reclassification rate.

In conclusion, we have shown that a systematic bulk reanalysis of class 3 CNLs identified in a diagnostic laboratory provided new diagnoses to some patients.

## Data Availability

All data produced in the present study are available upon reasonable request to the authors

