## Supplementary figures and images for "A SYSTEMATIC RE-ANALYSIS OF COPY NUMBER LOSSES OF UNCERTAIN CLINICAL SIGNIFICANCE"

### Supplementary Figure

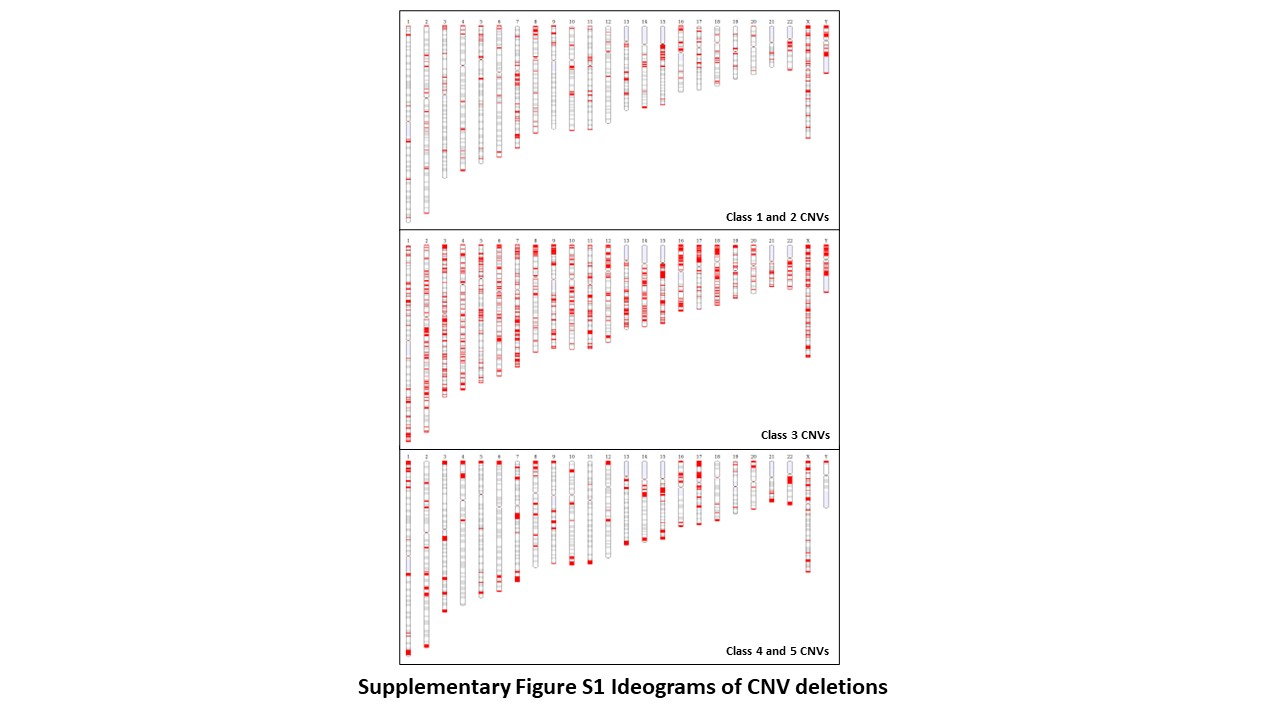
