## Supplementary Material for "A SYSTEMATIC RE-ANALYSIS OF COPY NUMBER LOSSES OF UNCERTAIN CLINICAL SIGNIFICANCE"

**Supplementary Table S1 Haploinsufficiency scores and genes according to CNL category**

| | Number of genes with available scores | | | Predicted Haploinsufficient genes (%) | | |
|---|---|---|---|---|---|---|
| **CNL class (Total no of genes)** | **pLI** | **RVIS** | **Decipher HI** | **pLI (%)** | **RVIS (%)** | **Decipher HI (%)** |
| **Pathogenic (2,611)** | 2,396 | 2,338 | 2,554 | 464 (19.37) | 493 (21.09) | 1,403 (54.93) |
| **Class 3 (2,543)** | 2,287 | 2,300 | 2,481 | 430 (18.80) | 559 (24.30) | 1,257 (50.67) |
| **Benign (433)** | 274 | 251 | 409 | 34 (12.41) | 40 (15.94) | 322 (78.73) |

**Supplementary Table S2: List of the prioritised 125 genes that have undergone scientist review**

| Gene | OMIM ID (*) | Number of cases | pLI ≥0.9 (Yes/No) | RVIS ≤ 0.01 (Yes/No) | DDG2P consequence | OMIM Phenotype # (Inheritance pattern) |
|---|---|---|---|---|---|---|
| ***NAA15*** | 608000 | 9[$] | Yes | Yes | LoF | 617787 (AD) |
| *MBD5* | 611472 | 6 | N/A | Yes | LoF | 156200 (AD) |
| *PIEZO2* | 613629 | 4 | No | Yes | LoF | 114300 (AD);108145(AD); 617146 (AR), 248700 (AD) |
| *TP63* | 603273 | 3 | Yes | Yes | LoF; Uncertain | 103285 (AD); 604292 (AD); 106260 (AD); 603543 (AD); 129400 (AD); 129400 (AD); 605289 (AD) |
| *SIX1* | 601205 | 3 | Yes | No | LoF; all missense/in frame | 608389 (AD); 605192 (AD) |
| *ZIC2* | 603073 | 3 | N/A | N/A | LoF | 609637 (AD) |
| *FOXG1* | 164874 | 3 | N/A | N/A | LoF | 613454 (AD) |
| *FREM1* | 608944 | 3 | N/A | N/A | LoF | 608980; 248450 (AR); 614485 (AD) |
| *NF1* | 613113 | 3 | Yes | Yes | LoF | 607785 (AD); 162210 (AD); 162200 (AD); 601321 (AD); 193520 (AD) |
| ***ZMYND11*** | 608668 | 3 | Yes | Yes | LoF | 616083 (AD) |
| *TBX1* | 602054 | 3 | Yes | No | LoF | 217095 ; 188400 (AD); 187500 (AD); 192430 (AD) |
| *SLC2A2* | 138160 | 3 | No | No | LoF | 125853 (AD); 227810 (AR) |
| *MITF* | 156845 | 2 | Yes | Yes | LoF; Uncertain | 617306 (AR); 614456; 103500 (AD); 193510 (AD); 103470 (AD) |
| *CDH23* | 605516 | 2 | No | Yes | LoF; all missense/in frame | 601386 (AR); 617540 (AD); 601067 (AR/DR); 601067 (AR/DR) |
| *MYH8* | 160741 | 2 | No | No | LoF; all missense/in frame | 608837; 158300 (AD) |
| *RERE* | 605226 | 2 | Yes | Yes | LoF | 616975 (AD) |
| *AUTS2* | 607270 | 2 | Yes | Yes | LoF | 615834 (AD) |
| *SATB2* | 608148 | 2 | Yes | Yes | LoF | 612313 (AD) |
| *ERCC6* | 609413 | 2 | No | Yes | LoF | 214150 (AR);133540 (AR); 278800 (AR); 211980 (AR); 613761 ; 616946 (AD); 600630 (AR) |
| *FOXP2* | 605317 | 2 | N/A | Yes | LoF | 602081 (AD) |
| *ATR* | 601215 | 2 | No | Yes | LoF | 614564 (AD); 210600 (AR) |
| *ABCC6* | 603234 | 2 | No | Yes | LoF | 614473 (AR); 264800 (AR); 177850 (AD) |
| *MYH11* | 160745 | 2 | Yes | Yes | LoF | 132900 (AD) |
| *ANKRD11* | 611192 | 2 | Yes | Yes | LoF | 148050 (AD) |
| *PIEZO1* | 611184 | 2 | No | No | LoF | 194380 (AD); 616843 (AR) |
| GNAS | 139320 | 2 | Yes | Yes | Activating; LoF | 219080; 174800; 166350 (AD); 617686; 103580 (AD); 603233 (AD); 612462 (AD); 612463 (AD) |
| ***CUX1*** | 116896 | 2 | Yes | Yes | N/A | 618330 (AD) |
| *MYH9* | 160775 | 1 | Yes | Yes | LoF; Uncertain | 603622 (AD); 153650 (AD); 153640 (AD); 600208 (AD); 155100 (AD); 605249 (AD) |
| *CDKN1C* | 600856 | 1 | No | No | LoF; GoF | 130650 (AD); 614732 (AD) |
| *ITPR1* | 147265 | 1 | Yes | Yes | LoF; DN; LoF; All missense/in frame | 206700 (AR, AD); 606658 (AD); 117360 (AD) |

| | | | | | | |
|---|---|---|---|---|---|---|
| *GJA1* | 121014 | 1 | No | No | LoF; All missense/in frame; Uncertain | 600309 (AD); 218400 (AR); 617525 (AD), 241550 (AR),164200 (AD); 257850 (AR); 104100 (AD); 186100 (AD) |
| *KIF1A* | 601255 | 1 | N/A | Yes | LoF; All missense/in frame | 614255 (AD); 614213 (AR); 610357 (AR) |
| *PLCG2* | 600220 | 1 | Yes | Yes | LoF; All missense/in frame | 614878 (AD); 614468 (AD) |
| *ZFPM2* | 603693 | 1 | Yes | Yes | LoF; All missense/in frame | 610187; 187500 (AD); 616067 (AD) |
| *TBC1D24* | 613577 | 1 | No | No | LoF; All missense/in frame | 220500 (AR); 614617 (AR); 616044 (AD); 615338 (AR); 605021 (AR) |
| ***ACTB*** | 102630 | 1 | Yes | No | LoF; All missense/in frame | 243310 (AD); 607371 (AD) |
| ***KMT2C*** | 606833 | 1 | Yes | N/A | LoF | 617768 (AD) |
| *EHMT1* | 607001 | 1 | Yes | Yes | LoF | 610253 (AD) |
| *CTNNB1* | 116806 | 1 | N/A | Yes | LoF | 114500;617572 (AD); 114550; 155255; 615075 (AD); 167000; 132600 |
| *TCF4* | 602272 | 1 | Yes | Yes | LoF | 613267 (AD); 610954 (AD) |
| *DSTYK* | 612666 | 1 | No | Yes | LoF | 610805 (AD); 270750 (AR) |
| *TGFB2* | 190220 | 1 | Yes | Yes | LoF | 614816 (AD) |
| *LRP4* | 604270 | 1 | No | Yes | LoF | 212780 (AR); 616304 (AR); 614305 (AR/AD) |
| ***MYT1L*** | 613084 | 1 | Yes | Yes | LoF | 616521 (AD) |
| *PARN* | 604212 | 1 | No | Yes | LoF | 616353 (AR); 616371 (AD) |
| *POU1F1* | 173110 | 1 | No | Yes | LoF | 613038 (AR/AD) |
| *DSG1* | 125670 | 1 | Yes | Yes | LoF | 615508 (AR); 148700 (AD) |
| *IGF1R* | 147370 | 1 | No | Yes | LoF | 270450 (AR/AD) |
| *TTN* | 188840 | 1 | No | Yes | LoF | 604145; 613765 (AD); 608807 (AR); 603689; 611705 (AR); 600334 (AD) |
| *RYR1* | 180901 | 1 | No | Yes | LoF | 117000 (AR/AD); 145600 (AD); 145600 (AD); 255320 (AR); 117000 (AR/AD) |
| ***PSMD12*** | 604450 | 1 | Yes | Yes | LoF | 617516 (AD) |
| ***BPTF*** | 601819 | 1 | Yes | Yes | LoF | 617755 (AD) |
| *SYNGAP1* | 603384 | 1 | Yes | Yes | LoF | 612621 (AD) |
| *LTBP3* | 602090 | 1 | Yes | Yes | LoF | 601216 (AR); 617809 (AD) |
| *CHD2* | 602119 | 1 | Yes | Yes | LoF | 615369 (AD) |
| *RELN* | 600514 | 1 | Yes | Yes | LoF | 616436 (AD); 257320 (AR) |
| *EPHB4* | 600011 | 1 | Yes | Yes | LoF | 617300 (AD) |
| *LEMD3* | 607844 | 1 | Yes | Yes | LoF | 166700 (AD); 166700 (AD) |
| *SOX5* | 604975 | 1 | Yes | Yes | LoF | 616803 (AD) |
| *EXT2* | 608210 | 1 | No | No | LoF | 133701 (AD); 616682 (AR) |
| *TWIST1* | 601622 | 1 | N/A | No | LoF | 123100 (AD); 180750 (AD); 101400 (AD); 617746 (AD) |
| *MEF2C* | 600662 | 1 | N/A | No | LoF | 613443 (AD); 613443 (AD) |

| | | | | | | |
|---|---|---|---|---|---|---|
| *EVC* | 604831 | 1 | No | No | LoF | 225500 (AR); 193530 (AD) |
| *EVC2* | 607261 | 1 | No | No | LoF | 225500 (AR); 193530 (AD) |
| *TGIF1* | 602630 | 1 | N/A | No | LoF | 142946 (AD) |
| *LMX1B* | 602575 | 1 | No | No | LoF | 161200 (AD) |
| *CRB1* | 604210 | 1 | No | No | LoF | 613835;172870 (AD); 600105 (AR) |
| *ATP8B1* | 602397 | 1 | No | No | LoF | 243300 (AR); 147480 (AD); 211600 (AR) |
| *NR2F2* | 107773 | 1 | N/A | No | LoF | 615779 (AD) |
| *CD96* | 606037 | 1 | N/A | No | LoF | 211750 (AD) |
| *TGFBR2* | 190182 | 1 | No | No | LoF | 614331; 133239; 610168 (AD) |
| *TNFRSF13B* | 604907 | 1 | No | No | LoF | 240500 (AR/AD); 609529 |
| *GATA4* | 600576 | 1 | No | No | LoF | 607941 (AD); 614430 (AD);615542 (AD);187500 (AD); 614429 (AD) |
| *SMAD6* | 602931 | 1 | No | No | LoF | 614823 (AD); 617439 (AD) |
| *SMAD3* | 603109 | 1 | No | No | LoF | 613795 (AD) |
| *SIX3* | 603714 | 1 | No | No | LoF | 157170 (AD); 269160 |
| *PTHLH* | 168470 | 1 | Yes | No | Increased gene dosage; LoF | 613382 (AD) |
| *NRXN2* | 600566 | 1 | Yes | Yes | LoF | N/A |
| ***NFIB*** | 600728 | 3 | Yes | Yes | LoF | N/A |
| *CTNNA2* | 114025 | 1 | Yes | Yes | LoF | N/A |
| ***PHF21A*** | 608325 | 1 | Yes | Yes | LoF | N/A |
| *OTUD7A* | 612024 | 8 | Yes | No | LoF | N/A |
| *NRXN3* | 600567 | 2 | Yes | Yes | LoF | N/A |
| *RYR3* | 180903 | 1 | Yes | Yes | LoF | N/A |
| *CACNA1C* | 114205 | 7 | Yes | Yes | Activating | 611875; 601005 (AD) |
| *NR3C2* | 600983 | 4 | Yes | Yes | N/A | 605115 (AD); 177735 (AD) |
| ***NTRK2*** | 600456 | 3 | Yes | Yes | All missense/in frame | 617830 (AD); 613886 (AD) |
| *RANBP2* | 601181 | 3 | Yes | Yes | All missense/in frame | 608033 (AD) |
| *ERBB4* | 600543 | 3 | Yes | Yes | N/A | 615515 (AD) |
| *ABL1* | 189980 | 2 | Yes | Yes | Activating | 617602 (AD) |
| ***CSNK2A1*** | 115440 | 2 | Yes | Yes | Activating | 617062 (AD) |
| *RYR2* | 180902 | 2 | Yes | Yes | N/A | 600996 (AD); 604772 (AD) |
| *PRKAG2* | 602743 | 2 | Yes | Yes | N/A | 600858 (AD); 261740 (AD); 194200 (?AD) |
| *BMPR2* | 600799 | 2 | Yes | Yes | N/A | 178600 (AD);178600 (AD); 265450 (AD) |
| *CFH* | 134370 | 2 | Yes | Yes | N/A | 126700 (AD); 609814 (AR/AD); 235400 (AR/AD); 610698 |

| | | | | | | |
|---|---|---|---|---|---|---|
| *CACNA1B* | 601012 | 2 | Yes | Yes | N/A | 614860 (AD) |
| *AKAP10* | 604694 | 2 | Yes | Yes | N/A | 115080 (AD) |
| *COL4A1* | 120130 | 1 | Yes | Yes | DN | 611773 (AD); 607595 (AD); 614519; 175780 (AD);180000 (AD);269160 |
| ***KIF2A*** | 602591 | 1 | Yes | Yes | DN | 615411 (AD) |
| *COL11A2* | 120290 | 1 | Yes | Yes | All missense/in frame; DN | 601868 (AD); 609706 (AR);614524 (AR)/AD); 184840 (AD); 215150 (AR) |
| *SMARCA2* | 600014 | 1 | Yes | Yes | All missense/in frame | 601358 (AD) |
| ***HECW2*** | 617245 | 1 | Yes | Yes | All missense/in frame | 617268 (AD) |
| *NEDD4L* | 606384 | 1 | Yes | Yes | All missense/in frame | 617201 (AD) |
| *RBPJ* | 147183 | 1 | Yes | Yes | All missense/in frame | 614814 (AD) |
| *KIF5C* | 604593 | 1 | Yes | Yes | All missense/in frame | 615282 (AD) |
| *DYNC1H1* | 600112 | 1 | Yes | Yes | All missense/in frame | 614228 (AD);614563 (AD); 158600 (AD) |
| *AKT3* | 611223 | 1 | Yes | Yes | All missense/in frame | 615937 (AD) |
| *PACS1* | 607492 | 1 | Yes | Yes | Activating | 615009 (AD) |
| *IGF2BP2* | 608289 | 1 | Yes | Yes | N/A | 125853 (AD) |
| *ADAM10* | 602192 | 1 | Yes | Yes | N/A | 615590; 615537 (AD) |
| *F2* | 176930 | 1 | Yes | Yes | N/A | 613679 (AR); 613679 (AR); 614390 (AD); 601367 ; 188050 (AD) |
| *DPP6* | 126141 | 1 | Yes | Yes | N/A | 616311; 612956 (AD) |
| *KCNH2* | 152427 | 1 | Yes | Yes | N/A | 613688 (AD); 613688 (AD); 609620 |
| *DNM2* | 602378 | 1 | Yes | Yes | N/A | 160150 (AD); 606482 (AD); 606482 (AD); 615368 (AR) |
| *ETV6* | 600618 | 1 | Yes | Yes | N/A | 601626; 616216 (AD) |
| *ACTN4* | 604638 | 1 | Yes | Yes | N/A | 603278 (AD) |
| *SLITRK1* | 609678 | 1 | Yes | Yes | N/A | 137580 (AD), 613229 (AD) |
| *LMNB2* | 150341 | 1 | Yes | Yes | N/A | 616540 (AR), 608709 (AD) |
| *STK11* | 602216 | 1 | Yes | Yes | N/A | 175200 (AD); 273300 |
| *MEN1* | 613733 | 1 | Yes | Yes | N/A | 131100 (AD) |
| *JPH3* | 605268 | 1 | Yes | Yes | N/A | 606438 (AD) |
| *SNRNP200* | 601664 | 1 | Yes | Yes | N/A | 610359 (AD) |
| *FLCN* | 607273 | 1 | Yes | Yes | N/A | 135150 (AD); 114500; 173600 (AD); 144700 |
| *WNK1* | 605232 | 1 | Yes | Yes | N/A | 201300 (AR); 614492 (AD) |
| ***MAPK8IP3*** | 605431 | 1 | Yes | Yes | N/A | 618433 (AD) |

Above is a list of 123 genes that were prioritised for scientist review (Please refer to **Error! Reference source not found.**)
Genes in **blue bold** font are those that were selected to undergo full CNV reclassification (**Error! Reference source not found.**)
$ 8 out of the 9 cases with *NAA15* loss were deemed artefactual on aCGH data inspection
